# Leveraging social network topology could improve the efficiency of SARS-CoV-2 epidemic control strategies in resource-limited contexts

**DOI:** 10.1101/2022.05.20.22275359

**Authors:** MV Evans, T Ramiadantsoa, K Kauffman, J Moody, C Nunn, JY Rabezara, P Raharimalala, TM Randriamoria, V Soarimalala, G Titcomb, A Garchitorena, B Roche

## Abstract

Targeted surveillance allows public health authorities to implement testing and isolation strategies when diagnostic resources are limited. When transmission patterns are determined by social contact rates, the consideration of social network topologies in testing schemes is one avenue for targeted surveillance, specifically by prioritizing those individuals likely to contribute disproportionately to onward transmission. Yet, it remains unclear how to implement such surveillance and control when network data is unavailable, as is often the case in resource-limited settings. We evaluated the efficiency of a testing strategy that targeted individuals based on their degree centrality on a social network compared to a random testing strategy in the context of low testing capacity. We simulated SARS-CoV-2 dynamics on two contact networks from rural Madagascar and measured the epidemic duration, infection burden, and tests needed to end the epidemics. In addition, we examined the robustness of this approach when individuals’ true degree centralities were unknown and were instead estimated via readily-available socio-demographic variables (age, gender, marital status, educational attainment, and household size). Targeted testing reduced the infection burden by between 5 - 50% at low testing capacities, while requiring up to 28% fewer tests than random testing. Further, targeted tested remained more efficient when the true network topology was unknown and prioritization was based on socio-demographic characteristics, demonstrating the feasibility of this approach under realistic conditions. Incorporating social network topology into epidemic control strategies is an effective public health strategy for health systems suffering from low testing capacity and can be implemented via socio-demographic proxies when social networks are unknown.

*French abstract available in Supplemental Materials

## INTRODUCTION

A key process of epidemic control is surveillance, whereby health systems test and isolate infectious individuals (1). However, many health systems lack the resources to test all symptomatic individuals and must allocate resources accordingly. This is particularly the case for emerging infectious diseases, such as SARS-CoV-2, where testing resources are unequally distributed across countries (2). As early as April 2020, the Africa Centers for Disease Control and Prevention called attention to the lack of SARS-CoV-2 diagnostics in the region (3). Indeed, serological surveys from multiple countries revealed infection burdens much higher than those predicted from case-based surveillance, an indication of underdiagnosis of SARS-CoV-2 cases by diagnostic systems (4–6). Testing is an integral part of the COVID-19 responses of sub-Saharan African countries (7), and the lack of adequate testing capacity is an impediment to public health efforts.

One way to mediate the limitation of low testing capacity is by using prioritized testing schemes, such as schemes that prioritize testing of only symptomatic individuals or testing of close contacts of known cases (8). Modeling studies have explored isolating sub-groups on social networks (9), prioritizing testing of close-contacts of infected individuals (10), expanding contact tracing to contacts of contacts (11), and reducing the overall number of contacts between individuals (12) as means to reduce transmission. Another potential strategy is to target surveillance of individuals or households based on their network characteristics, specifically their connectedness (13). For SARS-CoV-2, social contact heterogeneity has been identified as a primary driver of the distribution pattern of secondary infections, with a small proportion of infections causing a disproportionately high number of secondary infections (11,14). Given the strong role of social contacts, public health interventions that account for heterogeneity in social networks represent a promising avenue for implementing epidemiological surveillance in resource-limited contexts.

Targeted interventions can be particularly useful in contexts where surveillance and testing resources are limited, specifically by allocating these resources to individuals that may disproportionately contribute to community spread. A challenge to implementing this approach is that public health authorities rarely have access to social network data that would guide targeted surveillance (15,16). However, some socio-demographic variables predict connectivity on social networks and may therefore be useful proxies for the risk of spreading due to high contact rates when true social networks are not available. For example, a range of socio-demographic variables including age, household size and structure, income, and educational enrollment were used to predict age-specific contact rates across 152 countries (17). Similarly, socio-demographic variables such as gender, age, income, and education are used in marketing analytics to predict “central clients” (18). Thus, when it can be shown that socio-demographic variables predict network centrality, it may be possible to use those variables as proxies for the risk of onward transmission in targeted surveillance approaches. This would greatly increase the feasibility of including network topology in epidemic control.

Here, we use epidemic simulations on empirical and simulated social networks to investigate the effectiveness of targeted testing of highly connected individuals to control an epidemic when testing capacity and social network information is limited. We simulate SARS-CoV-2 outbreaks on two close-contact social networks derived from social and spatial movement data on individuals living in rural communities in the Sambava district of the SAVA region in northeastern Madagascar (19). We then compare the effectiveness of testing strategies that target testing based on social connectivity to those that test randomly using the full knowledge of the social network, evaluating the time needed to control the epidemic, the total infection burden, and the number of tests needed. Finally, we repeat the simulations using socio-demographic variables (age, household size, marital status, educational attainment) from the study population to guide targeted testing, rather than information on individuals’ degree centralities from the social networks, thus investigating whether these commonly available data are suitable proxies for heterogeneity in transmission.

## RESULTS

### Madagascar Social Network Topology

The social networks from Mandena and Sarahandrano in rural Madagascar had characteristics of networks where heterogeneous transmission patterns are likely to occur. The Mandena network contained 120 nodes and 4136 total edges, with a mean node degree of 34.47 and normal degree distribution. However, the distribution of edge weights was strongly left-skewed. The mean edge-weight was 0.18, but less than two percent of edges had a weight above 1 (equal to 24 hours over a week-long period). The majority of edge weights were below 0.05, equivalent to 80 minutes of close contact a week. The Sarahandrano network contained 318 nodes and 16140 total edges, with a mean node degree of 50.7 and a normal degree distribution. Like the Mandena network, the distribution of edge weights was strongly left-skewed, but degree centrality was overall higher than on the Mandena network. The mean edge-weight was 0.31, with 614 edges, or 3.8%, having a weight above 1. Fifty-percent of edge-weights were below 0.08.

### Control Efficiency

We compared the control efficiency of targeted and random surveillance strategies at low and high testing capacities, corresponding to the ability to test 25% and 100% of the population monthly. The targeted strategy had the largest effect on reducing daily incidence while using the fewest tests, particularly at a 25% testing capacity (Fig. 1, Fig. 2). For all three measures of effectiveness, targeted testing was more effective on the Mandena network than the Sarahandrano network (Fig. 2, Table 1). An uncontrolled epidemic, with no testing, resulted in a median of 0.82 infections per capita on the Mandena network and 0.94 infections per capita on the Sarahandrano network (Table 1, Fig. 2). In contrast, at a monthly testing capacity of 25%, targeting highly connected individuals resulted in a median of 0.41 and 0.90 new infections per capita on the Mandena and Sarahandrano networks, respectively (Table 1, Fig. 2). The infection burden decreased with increasing testing capacity for both control strategies, but the random testing strategy was most sensitive to an increase in testing capacity (Table 1, Fig. 2).

**Figure 1.**
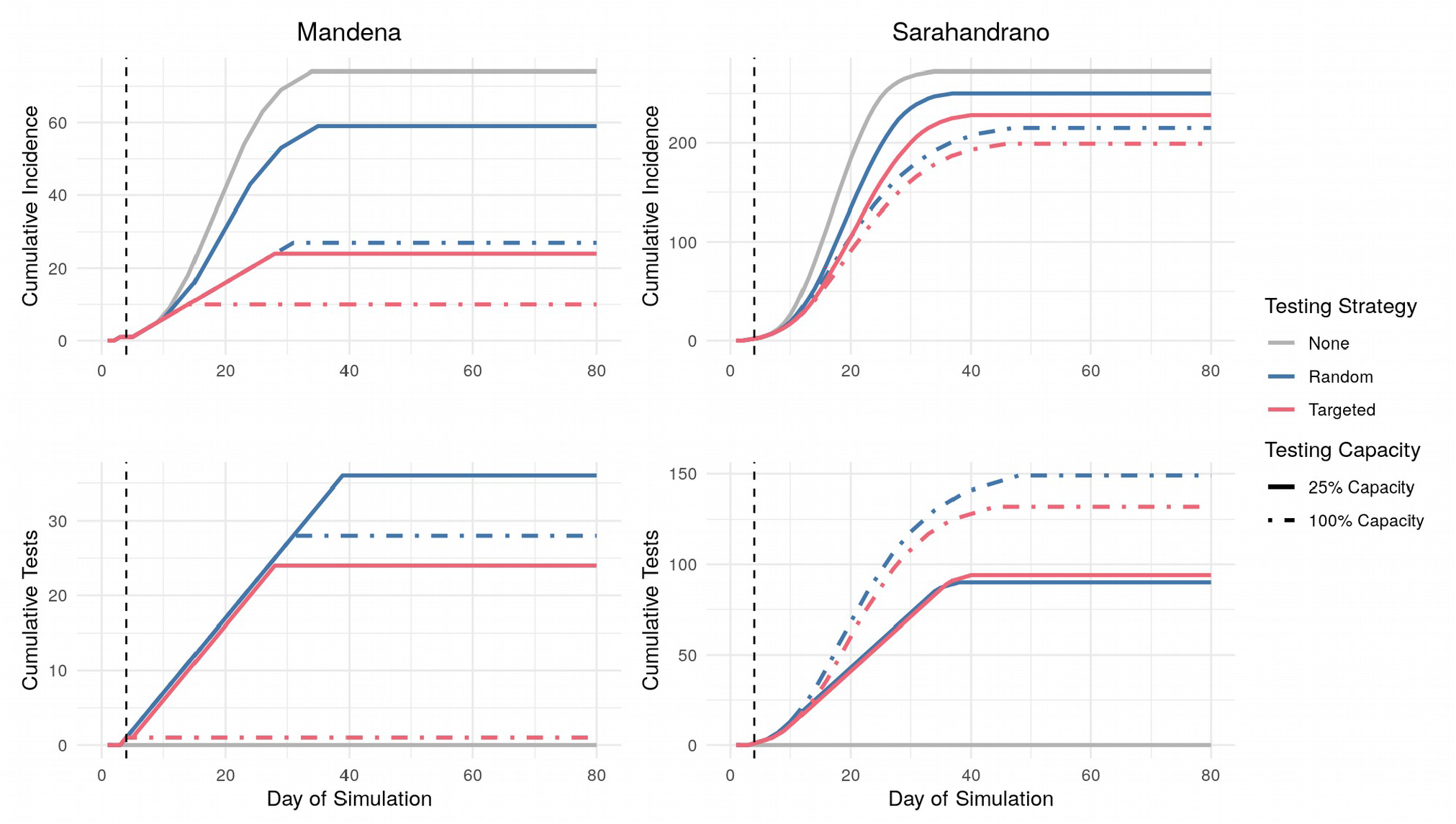
Targeted testing reduces daily incidence while requiring fewer tests than random testing. Cumulative daily incidence (top row) and cumulative tests required (bottom row) for the two testing strategies across two testing capacities on the Mandena and Sarahandrano networks. Testing capacities refer to ability to test a percentage of the total population monthly. The vertical dashed line represents the start of the control strategies at day 4. Lines represent median values from 1000 simulations.

**Figure 2.**
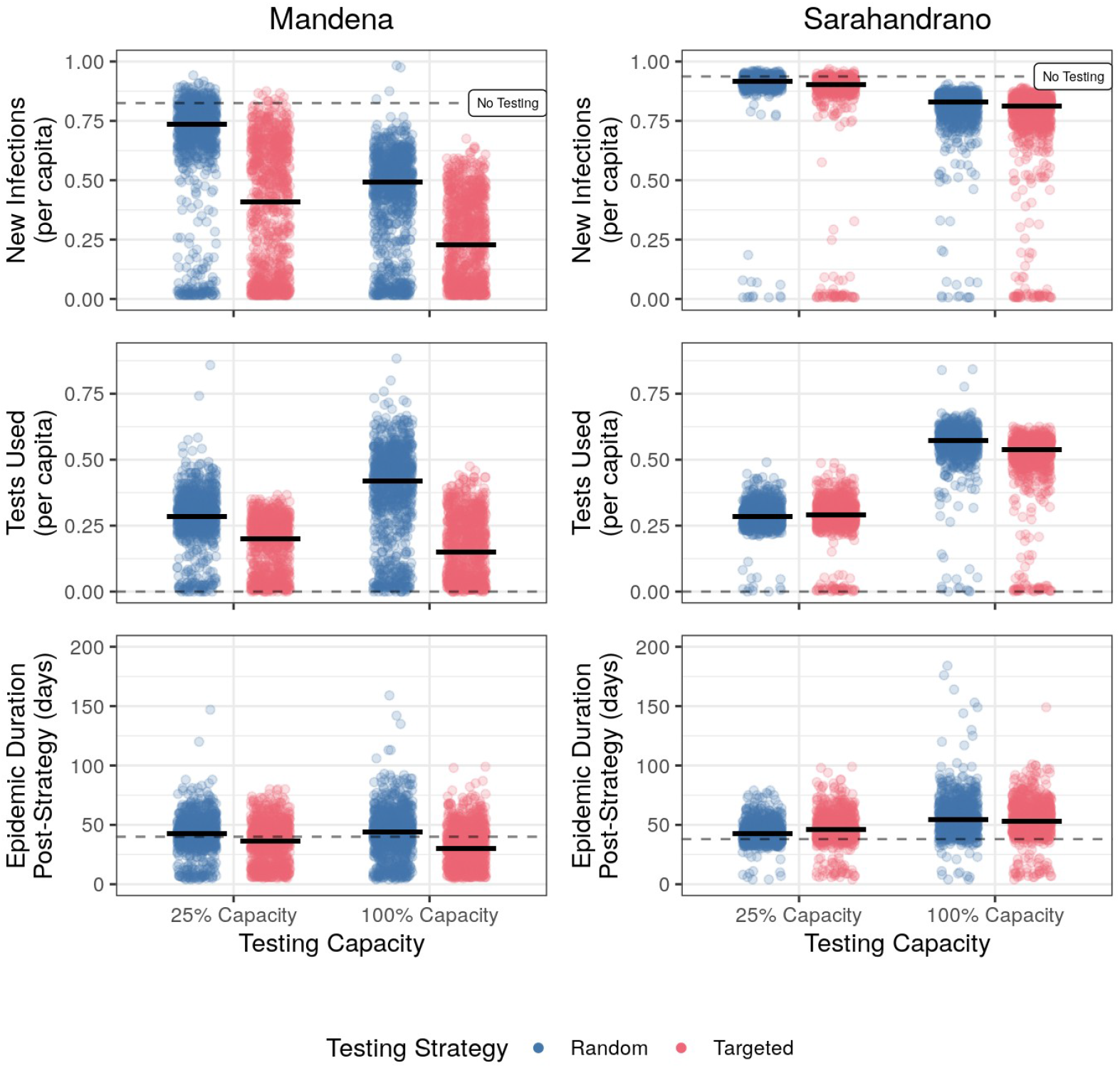
Targeted testing reduces the total infection burden and the number of tests needed, even at low testing capacities. Comparison of efficiency of two control strategies at two testing capacities on the Mandena and Sarahandrano networks. Testing capacities correspond to monthly testing capacities equal to testing 25% and 100% of the total population. The dashed black line represents median values from simulations with no testing. Raw data is represented by points and median values per strategy are represented by bold horizontal lines. The figure displays results from 1000 simulations for each combination of testing capacity and control strategy.

**Table 1.** Median and 95% CI of efficiency metrics for two control strategies on two empirical social contact networks from rural Madagascar. Represents median and confidence intervals from 1000 simulations. Testing capacity corresponds to monthly testing capacity, with 100% equal to the ability to test the full population monthly. Note that efficiency at 0% testing capacity is the same for both strategies because it represents the control strategy of no testing.

|  |  | Mandena Network |  | Sarahandrano Network |  |
| --- | --- | --- | --- | --- | --- |
| Efficiency Metric | Testing Capacity | Random Testing | Targeted Testing | Random Testing | Targeted Testing |
| Epidemic Duration | 0% | 44<br>(13,67) | 44<br>(13,67) | 42<br>(33,58) | 42<br>(33,58) |
|  | 25% | 46<br>(11,73) | 40<br>(10.75, 71) | 46<br>(36,67.1) | 50<br>(16,76) |
|  | 100% | 48<br>(11,88) | 34<br>(11,71) | 58<br>(39,91) | 57<br>(16,89) |
| Infections Per Capita | 0% | 0.82<br>(0.03,0.87) | 0.82<br>(0.03,0.87) | 0.94<br>(0.91,0.97) | 0.94<br>(0.91,0.97) |
|  | 25% | 0.73<br>(0.02,0.87) | 0.41<br>(0.02,0.78) | 0.92<br>(0.87,0.95) | 0.90<br>(0.01,0.94) |
|  | 100% | 0.49<br>(0.02,0.69) | 0.22<br>(0.02,0.57) | 0.83<br>(0.54,0.88) | 0.81<br>(0.01,0.87) |
| Tests per Capita | 25% | 0.28<br>(0,0.45) | 0.20<br>(0.01,0.32) | 0.28<br>(0.22,0.38) | 0.29<br>(0.01,0.41) |
|  | 100% | 0.42<br>(0.02,0.64) | 0.15<br>(0.01,0.39) | 0.57<br>(0.39,0.66) | 0.54<br>(0.01,0.6) |

Testing was more efficient when targeting highly connected individuals on the Mandena network, requiring less than three-quarters of the number of tests needed when testing randomly at 25% capacity. This was equivalent to 34 tests when testing randomly and 24 tests with targeted testing. In contrast, both testing strategies used similar numbers of test on the Sarahandrano network, approximately 89 tests. Targeted testing only shortened the epidemic length on the Mandena network (Table 1, Fig. 2), where it was able to stop transmission chains earlier in the epidemic than random testing (Fig. 1, Fig. 3). On the Sarahandrano network, both control strategies flattened the epidemic curve by reducing the number of infections, and this prolonged the epidemic by preventing rapid spread through the population, as was seen in the simulations without testing (Fig. 1, Fig. 2).

**Figure 3.**
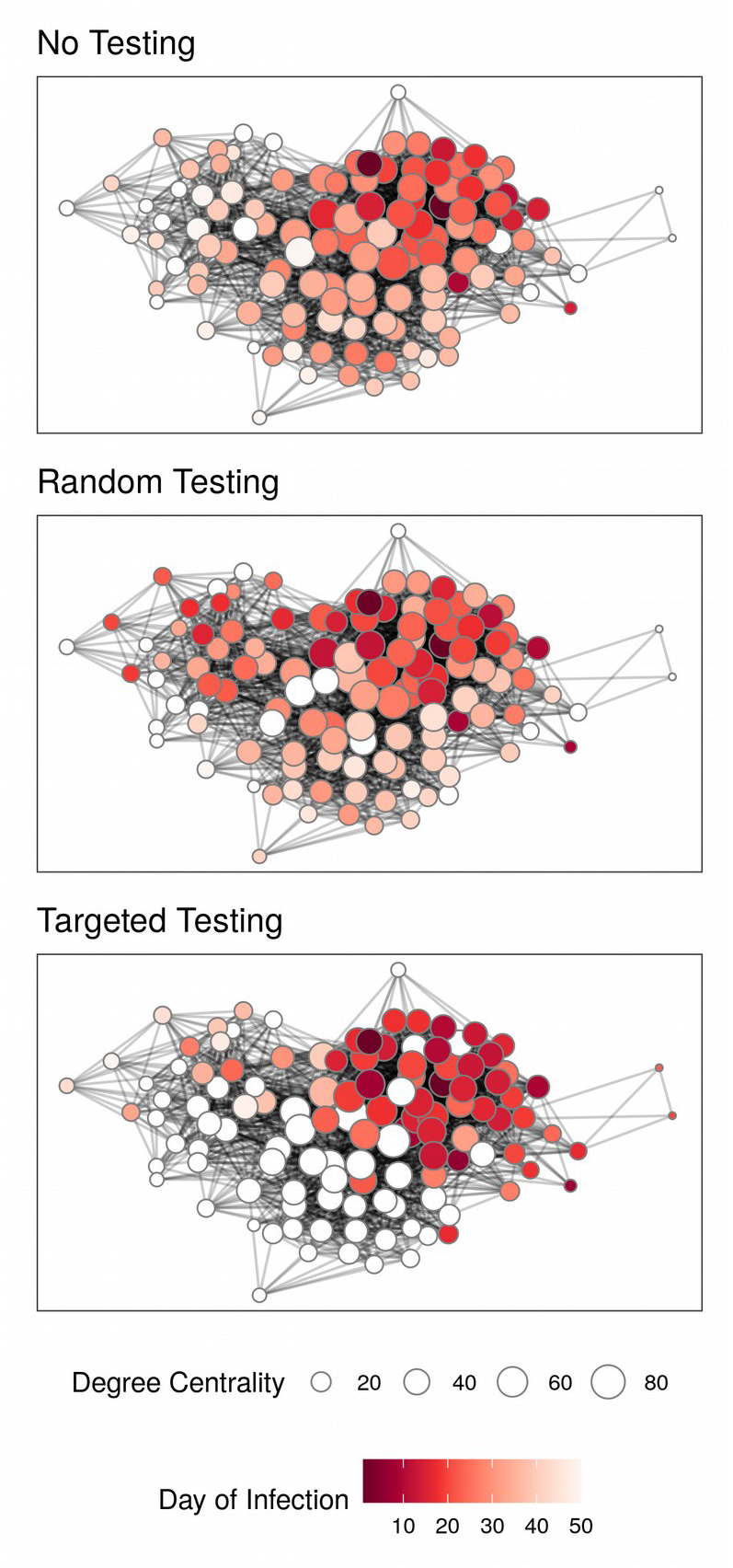
Targeted testing most efficiently reduces disease spread across social networks. Final epidemic spread of one simulation on the Mandena network for three different control strategies. Nodes are represented by points, colored based on day of infection, and sized according to their degree centrality. Nodes that were never infected are white. All control strategies used a monthly testing capacity of 25%.

Visualizing a simulated SEIR epidemic on the Mandena network illustrates how each strategy works at a fine scale (Fig. 3). The strategy of no control allowed for the highest infection burden, including individuals with low centrality who were infected later in the epidemic than in other strategies. Testing randomly did stop some transmission chains, but transmission rapidly spread early in the epidemic, with the highest daily incidence of 7 cases on day 32 of the epidemic. In contrast, targeted testing slowed transmission by halting transmission chains that would result in a high number of secondary infections (Fig. 3); daily incidence never rose above 5 cases. This resulted in a longer epidemic duration of 50 days, but daily incidence during the second half of the epidemic was never above one new infection per day.

We conducted sensitivity analyses to identify scenarios in which targeted testing is more efficient than random testing. Across all three categories of the sensitivity analyses, targeted testing remained the most efficient strategy (Fig. S4.1 - S4.5). The efficiencies of both strategies became similar when testing capacity was low and testing began 15 days after the start of the epidemic, after which most individuals in the network had already been infected, and testing could do little to control the infection burden (Fig. S4.1). The relative efficiency was also reduced at low levels of ascertainment, particularly below levels of 0.25, when infected individuals only had a 0.25 probability of being correctly identified and tested (Fig. S4.2). At this point, the low accuracy of ascertainment reduced the ability to identify highly connected individuals for targeted testing. Targeted testing was more effective than random testing at all transmission rates, but was most effective at transmission rates between 0.15 and 0.23 (Fig. S4.3). At lower transmission rates, daily incidence was low enough that random testing could test most infected individuals. At higher transmission rates, the epidemic spread quickly and many individuals were already infected when testing began on day 4. Higher testing capacities increased the relative effectiveness of targeted testing at high transmission rates and decreased the relative effectiveness at low transmission rates. Sensitivity analyses concerning network size were highly stochastic; however, targeted testing always resulted in a lower infection burden than random testing (Fig S4.4). Similarly, there were few clear differences between the two testing strategies on networks of different assortativity values, with very small magnitudes of difference between the two strategies (Fig. S4.5).

### Applying Control Strategies to Networks with Unknown Topologies

We focused on five socio-demographic variables as predictors of an individual’s degree centrality: age, gender, household size, marital status, and education level. A model including socio-demographic variables did a poor job of predicting degree percentile across the two networks (R^2^ = 0.03). However, the model was able to rank individuals by degree centrality (Mandena Spearman’s ρ = 0.15 (p=0.09); Sarahandrano Spearman’s ρ = 0.18 (p=0.002)). The model distinguished high-degree individuals from low-degree individuals: the top ten individuals in each network had a predicted degree that was on average approximately twice as high as the bottom ten individuals (Fig. S3.3). Marital status was the only variable included in all models within 4 AIC units of the top model, but all socio-demographic variables were included in the averaged model. Specifically, cohabiting individuals had lower degree centrality than single individuals. Further details are reported in the Supplementary Materials.

Despite the poor capacity of socio-demographic variables to predict degree percentile, the predictions based on socio-demographic variables performed remarkably well in guiding the targeted testing strategy. Indeed, testing based on these centrality “proxies” was as effective as using “true” centralities obtained from a social network (Fig. 4). The infection burden was slightly higher when using estimated degree centralities to target testing, but it remained lower than strategies of no testing and testing randomly. Although socio-demographic characteristics were not strong predictors of absolute degree centrality values, we found that their ability to differentiate between very high and low connected individuals was enough to successfully implement a control strategy that considers network topology.

**Figure 4.**
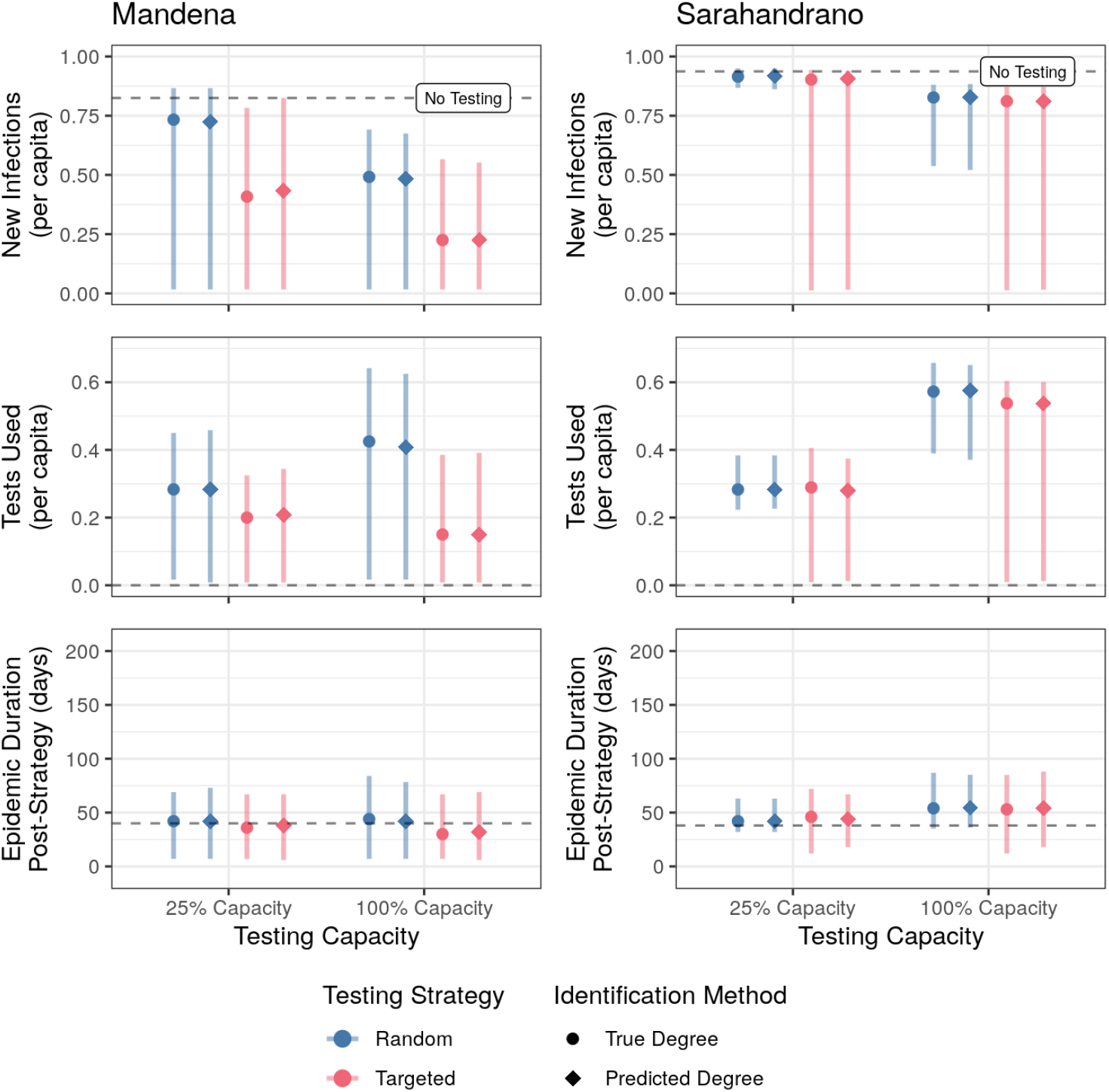
Targeted testing based on individuals’ degree centrality proxies performs similarly to targeted testing based on individuals’ known degrees. Comparison of efficiency of two control strategies at two testing capacities on the Mandena and Sarahandrano networks using true degree centralities (circles) or degree centralities predicted from socio-demographic variables (diamonds).Testing capacities correspond to monthly testing capacities equal to testing 25% and 100% of the total population. The dashed black line represents median values from simulations with no testing. Points represent the median and error bars the 95%CI based on 1000 simulations.

## DISCUSSION

In the face of global diagnostic and vaccine inequity, many countries are tasked with developing novel public health interventions that optimize limited diagnostic capacities to control the SARS-CoV-2 epidemic. Given the role of social contact variation in community transmission of SARS-CoV-2, we explored whether control strategies that consider social network topologies, specifically individuals’ degree centralities, could limit disease transmission while using fewer resources than current, non-prioritized testing strategies. We found that strategies that target well-connected, infected individuals are the most effective, reducing epidemic duration and overall infection burden, particularly at low testing capacities. In fact, in simulations on empirical social contact networks from rural Madagascar, targeted testing reduced the infection burden and shortened the epidemic even at a testing capacity of only one test per day, equivalent to a monthly testing capacity of 25% of the population. These strategies were robust even when targeting was imperfect due to unknown network topologies and based solely on socio-demographic variables. Importantly, this implies that this theoretical network-based approach is feasible in practice because it can be implemented using commonly available data on individuals, such as age, marital status, and household size. Our findings therefore demonstrate the benefits of considering social network topology in data-driven epidemic control strategies even when social network data is incomplete or not available.

We find that strategies that prioritize testing highly connected individuals offer the most benefit in contexts with low testing capacities. In our simulations, this is achieved by controlling the epidemic before it reaches the point at which limited testing capacity cannot contain it. However, even when the start of testing is delayed by 24 days, the targeted strategy can avoid on average nine infections on the Mandena network, or 0.075 infections per capita (Fig. S4.1). Early, aggressive testing has been used to successfully control SARS-CoV-2 in several countries (South Korea: (20), New Zealand: (21)), and a similar mechanism explains why strategies that target highly connected individuals are so efficient in our simulations. In addition to delayed testing, high transmission rates can result in epidemics that targeted testing is unable to control at limited testing capacities. This was seen in our sensitivity analyses (Fig. S4.3) and on the Sarahandrano network, where higher average edge weights resulted in higher community transmission than on the Mandena network. On the Sarahandrano network, 12 individuals (95% CI: 2-36) had already been exposed by the start of testing on day 4, and neither testing strategy effectively controlled an epidemic at low testing capacities (Fig. 2, Table 1). The relatively low connectivity between rural communities in sub-Saharan Africa has been proposed to slow the epidemic pace of SARS-CoV-2, as compared to the US or Europe (22). However, at the community-scale, such as those portrayed in our empirical networks, connectivity between individuals was high. Therefore, at this scale, epidemics may spread rapidly following an initial introduction, and implementing measures quickly are key to limiting infections. Notably, at the beginning of the pandemic in 2020, many African countries implemented control measures more quickly than European countries (23), effectively limiting spread during the first wave of the epidemic. Our simulations show that implementing targeted testing strategies at the beginning of local epidemics similarly reduces disease burdens while requiring few testing resources.

While the concept of developing a disease control strategy configured by a social network is not novel (12,24,25), this study is one of the few that explicitly considers limited testing capacities on par with those in low-income countries. Madagascar tested 26,425 individuals (less than 0.01% of the total population) for SARS-CoV-2 between March and September 2020 (26), only a fraction of the testing capacity required by mass-testing campaigns that have been implemented elsewhere (e.g. Slovakia, (27)). This is further complicated by the relative remoteness of some communities, with more than 50% of the population living further than two hours from a hospital (28). In both Mandena and Sarahandrano, for example, no SARS-CoV-2 testing has been available to date. While cost and physical access to testing are significant barriers to disease control in Madagascar (29), our findings suggest that, if and when testing is available to rural communities, targeted testing can mitigate the negative impact of limited testing capacity on epidemic control. For example, antigen-based rapid diagnostic testing is inexpensive, does not require refrigeration, provides instant results, and could be implemented at a local scale via outreach teams of skilled health workers (7). However, for prioritized testing schemes such as this to be possible, the global inequality in access to rapid diagnostic tests must first be overcome (30).

Many theoretical studies have shown the effectiveness of incorporating network topology into epidemic control strategies (31,32), but the feasibility of doing so has been questioned because the true social network is almost never known. One alternative is occupationally-targeted strategies that target high contact rates or high risk environments (e.g. health-care or food service worker) (33). However, in rural communities such as Mandena and Sarahandrano, all community members are agriculturalists, with little variation in occupation. To overcome this obstacle, we considered socio-demographic predictors of network centrality to guide targeted testing rather than the true values of network centrality or occupationally-based targeting. While demographic predictors did not accurately rank individuals by degree centrality, they were able to distinguish between individuals with high and low contact rates. Despite this imperfect predictive performance, estimated measures of degree centrality based on common socio-demographic variables performed as well as “true” degree centrality when used to prioritize testing schemes in our simulated epidemics. Health authorities can implement targeted control strategies by taking into account easy to measure individual characteristics (e.g. age, gender, household size, marital status), many of which are available in healthcare and governmental records, or can be quickly generated through surveys. The exact socio-demographic variables to include will vary depending on local demographics and cultural practices, and will require input from local experts. For example, in urban communities or communities with higher market integration, degree centrality may be more closely related to economic activities (e.g. workforce labor, commuting dynamics, income levels), rather than the social ties (e.g. marital status, household size) found on our rural networks. Further, when an incomplete network exists, it could be used to validate whether proposed demographic variables covary with degree centrality in that community. We found that even imperfect models can inform prioritization strategies if they are able to differentiate the most connected from the least connected individuals. This suggests that a high level of predictive performance is not necessary to successfully integrate social network topologies into control strategies via socio-demographic proxies. This robustness to low predictive performance of socio-demographic variables further supports the feasibility of this approach in settings where social network and socio-demographic data quality and availability may be low.

We included two social networks from rural Madagascar in our study to assess the generalizability of our results among rural communities. Targeted testing was more efficient on the Mandena network than the Sarahandrano network, where no testing strategy could control epidemics at low testing capacities. Epidemics spread very quickly on the Sarahandrano network due to the combination of highly connected individuals (mean degree centrality of 50.7) and a high disease transmission rate (R_0_ = 12.48). Our sensitivity analyses confirmed that the effectiveness of targeted testing was dependent on the transmission rate of the disease (Fig S4.3), and targeted testing may not be appropriate when disease transmission is extremely low or high. This agrees with other mathematical models of SARS-CoV-2, which show that the effectiveness of testing to control epidemics becomes limited at increasing transmission rates (10,13). However, socio-demographic variables performed equally well as proxies for true degree centrality on both networks (Fig. S3.3, Fig. 4). Therefore, although the benefit of targeted testing is dependent on the characteristics of the pathogen and social network, the ability to implement targeted testing via socio-demographic proxies appears generalizable, at least in rural contexts. Future research on a diversity of social networks is needed to explore the efficiency of these strategies in other contexts.

Our social networks represented realistic, but necessarily simplified versions, of true social networks. For example, we assumed social contacts were static and did not change, either as a result of dynamic social behavior (34), or changes in behavior due to the epidemic (e.g. social distancing). However, our simulations were relatively short, lasting less than three months on average, and static networks have been successfully used to model rapid epidemics, such as SARS-CoV-1 (35). Our social networks necessarily only included individuals over 18 years old, who represent less than 50% of the population nationally. Although contact patterns may differ for individuals under 18 years old, our sensitivity analyses found that the targeted strategy was most effective for a variety of network topologies, suggesting these results will hold when applied to the full network of all ages. In addition, the small size of our networks (120 and 318) meant that epidemics were short and highly influenced by stochasticity, which we attempted to control for by simulating 1000 epidemics with different initial conditions. By using empirical social contact networks, we included realistic social network topologies that more accurately represent exposure risk in rural Madagascar than simulated networks or networks based on studies from the Global North, where the majority of social networks originate from. A recent meta-analysis found only four social contact studies, less than 7% of those included in the meta-analysis, incorporated data from sub-Saharan Africa (36). Expanding social network data collection outside of the Global North would allow for more realistic and context-specific estimates of disease dynamics on social networks globally.

Incorporating social network topology into SARS-CoV-2 interventions greatly increases the efficiency of control strategies under limiting testing capacities. Our theoretical model found that prioritized testing of highly-connected individuals reduced the infection burden while using fewer tests than random testing under low-testing capacity. These control strategies were effective even when the true network topology was unknown and testing was prioritized using only common socio-demographic variables. Incorporating a similar strategy could aid countries with limited diagnostic resources to manage the SARS-CoV-2 epidemic and other infectious disease outbreaks within their communities. Importantly, this theoretical study focused on testing to control the epidemic at the population level, and assumed that diagnostics are not tied to treatment. SARS-CoV-2 control strategies should be adapted according to a country’s public health policy to ensure that vulnerable populations do not lose access to diagnostics and treatment. While our study focused on SARS-CoV-2, testing strategies that consider network topology may be useful for other diseases with similar transmission pathways, particularly when social contact drives transmission. This simulation approach could be easily re-parameterized to characterize different diseases of interest. As social contact network data becomes more widely available, considering social network topology is a promising method for allocating limited resources during public health crises.

## METHODS

### Social Contact Networks

We modeled SARS-CoV-2 dynamics using a susceptible-exposed-infected-recovered (SEIR) model simulated across undirected networks where each individual was represented explicitly as a node in the network and their contacts by edges. Importantly, we modeled epidemics on empirical social networks, rather than simulated ones, to ensure our results were most relevant to settings with limited testing capacities. Empirical contact networks contain unique network topologies that may be lacking from simulated networks and more accurately represent the true social structure of a population, with important consequences for disease transmission (37). However, half of all empirical social contact networks used in epidemic modeling are derived from European contexts (36), where SARS-CoV-2 testing capacities are relatively high. To ensure our results were most relevant to settings with limited testing capacities, we simulated epidemics on two contact networks obtained from rural communities in Madagascar, where testing capacities are currently limited.

The social contact networks were constructed using survey and GPS tracker data of consenting adults (over 18 years of age) living in two villages in the SAVA region of northeastern Madagascar. The first village, Mandena (14°28’36″ S 47°48’50″ E), as described by Kauffman et al. (19), has approximately 2700 people (based on census data from local authorities). The second village, Sarahandrano (14°36’27’’ S 49°38’50’’), is home to approximately 900 people. Data collection occurred over 7 weeks in Mandena and over 3 separate sampling periods, ranging from 5 to 8 weeks each over a 9-month time period, in Sarahandrano. Subjects participated in a social network survey that asked for information on individuals with whom they have regular contact or supportive social relationships. These social surveys were used for recruitment of additional individuals into the study via snowball sampling (38). Subjects (n = 123 and 321, respectively) were provided with a GPS tracker programmed to record a location every three minutes to wear for at least one week. Participants wore the GPS for at least one week, with a subset of individuals agreeing to wear a new, freshly charged GPS for additional weeks (n = 76; 61.8% and n=154; 48.0% for Mandena and Sarahandrano, respectively). The close-contact networks were then imputed to account for individuals wearing GPS trackers in different weeks of the study. Edges in the network represent the probability a dyad came into close contact and the predicted proportion of 3 minute-periods of close-contact between dyads over the number of possible contacts in one week (details in supplement). We included edges with a weight above a threshold value equal to at least one hour of contact during the week, resulting in final network sizes of 120 and 318 for the Mandena and Sarahandrano networks, respectively. To create a time-integrated network, we rescaled the weight so that a value of 1 corresponded to 24 hours in contact over the course of the week.

### Epidemic Model Simulations

At each time step, equivalent to one day, an individual could become susceptible, exposed, pre-symptomatic, infected (symptomatic and asymptomatic) or recovered (Fig. S2.1). For each contact event (e.g. edge between a susceptible and infected node), a susceptible individual’s probability of becoming exposed was a function of the transmission probability *β* of the infected contact and the edge weight. Therefore, a susceptible individual’s probability of becoming exposed at each time step was a function of how many infected contacts (*j)* they were connected to on the network and their transmission rates (*β*_*j*_) (Eq. 1) :

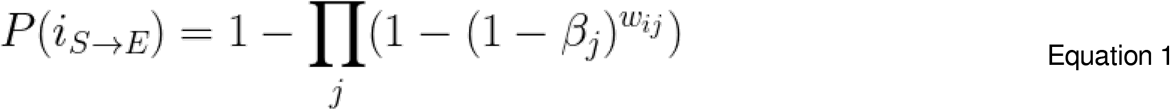

Where *i* is the susceptible individual, *j* is the infected individual connected to i, *β*_*j*_ is the time-dependent transmission rate of individual *j*, and *w*_*ij*_ is the edge-weight between nodes *i* and *j*. Individuals then moved through exposed, pre-symptomatic, infected, and recovered compartments following time-dependent transition rates.

Each transition event was drawn from a Bernoulli distribution defined by a daily transition probability rate. Time-dependent transition rates more accurately describe SARS-CoV-2 epidemic dynamics than memoryless transition rates, which assume transition rates are independent of the time spent in the compartment (39). Therefore, we relaxed the memoryless assumption of all transition rates, and parameterized the model to approximate an outbreak of SARS-CoV-2 Omicron variant. The incubation period (α) followed a gamma distribution with a mean of 3.1 days () followed a gamma distribution with a mean of 3.1 days (*sd* 2.6), including one day of pre-symptomatic transmission (40). The mean recovery period (γ) followed a) followed a Poisson distribution with a mean of 9.67 days (41) (Fig. S2.2). We modeled waning immunity (ζ) using) using a gamma distribution with a mean of 90 days (*sd* 20.12), parameterizing the scale of the distribution so that recovered individuals had a probability of becoming susceptible beginning 40 days post infection, with that probability increasing logistically until 150 days post-infection, when no immunity remained (Fig S2.2).

The transmission rate *β* was time-dependent: infected individuals had a transmission rate equal to *β* upon entering the infected class, and this transmission rate decreased exponentially as they spent more time infected (Fig. S2.3). *β* was parameterized to approximate realistic effective reproductive numbers at the beginning of the outbreak given the rapid susceptible depletion on smaller networks. This resulted in a mean estimated reproductive number of 8.99 on the Mandena network and 12.48 on the Sarahandrano network during the first 5 days (see supplement for further details), appropriate for a completely susceptible population exposed to the Omicron variant of SARS-CoV-2 (42). A range of transmission rates were explored via a sensitivity analysis reported below. The pre-symptomatic transmission rate was defined as 50% of the infectious transmission rate (43). We assumed thirty percent (30%) of infected individuals were asymptomatic (44), and that asymptomatic individuals had a transmission rate equal to 30% of the transmission rate (*β*) of symptomatic individuals during the pre-symptomatic and infectious stage (45). Further details on model specification are provided in the Supplemental Materials.

Each simulation was initiated by randomly selecting two individuals to be exposed. These exposed individuals thus started the simulation on the first day of their latent period. The number of susceptible, exposed, pre-symptomatic, infected (asymptomatic and symptomatic), isolated, and recovered individuals were recorded at each time step. Epidemics were simulated until no exposed, presymptomatic, infected, or isolated individuals remained, which we define as the full duration of the epidemic.

### Evaluating Control Strategies

We considered two different testing strategies. One strategy targeted testing based on social network connectivity. In this strategy, individuals were tested in order of descending degree centrality (e.g. the number of nodes they are connected to); hence, the most well-connected individuals were tested first. The other strategy tested individuals randomly, without consideration of network topology. We only considered passive surveillance, which tests infected, symptomatic individuals. Testing took place at the end of each simulated time-step (daily), after individuals had gone through transitions at that time step. To account for imperfect surveillance, symptomatic individuals that were identified to be tested had a 0.75 probability of being successfully contacted and tested. This parameter was explored further in the sensitivity analyses. Infected individuals that were positively identified via testing were isolated by moving them immediately to the isolated class. We did not include a delay between testing and isolation because we assumed diagnostics were limited to rapid antigen tests, which is more likely in Madagascar and other low-resource contexts given the limited resources for PCR diagnostics (29). Isolated individuals remained isolated until seven days post symptom onset, after which they moved to the recovered class.

Household transmission can reduce the effect of isolation on epidemic control by leading to imperfect isolation (46). Because the social networks did not include information on household membership, we approximated household transmission during isolation by allowing exposure events between an isolated, infected individual and susceptible contacts with an edge weight greater than 1 (equivalent to an average 24 hours together over one week). The transmission rate during these exposure events was scaled by a factor of 0.4 to account for other behaviors (distancing, mask wearing) that an isolated individual would be practicing while under isolation (47).

In addition to two testing strategies, we considered low and high testing capacities, corresponding to monthly testing capacities of 25% and 100% of the total population. Testing began on day four of all simulations, with a range of start dates explored in the sensitivity analyses. All strategies, including a control of no testing, were simulated 1000 times.

We evaluated each strategy and testing capacity combination based on how efficiently it controlled the epidemic, defined as the time and resources spent until no exposed, pre-symptomatic, infected, or isolated individuals remained. We used three metrics to evaluate the outcomes: the duration of the epidemic, the cumulative number of infected individuals per capita, and the number of tests used. We assessed each strategy and testing capacity based on its ability to reduce the infection burden and the length of the epidemic while minimizing the tests needed.

### Sensitivity Analysis

We assessed the robustness of our results by varying three categories of parameters in our simulations: intervention parameters (start date and imperfect surveillance rate), biological parameters (transmission rate), and network parameters (network size and assortativity). We compared the efficiency of the two different control strategies via the same measures used in the main analysis. Further details on these methods and results are reported in the Supplemental Materials.

### Applying Control Strategies to Networks with Unknown Topologies

To examine the applicability of these strategies to real-world scenarios, we compared the effectiveness of the two strategies when applied to a population where the true degree distribution is unknown and identification of individuals for targeted testing is derived from common socio-demographic variables. First, we fit a statistical model to predict each individual’s degree percentile in their respective network using the following socio-demographic variables: age, gender, household size, marital status (single vs. cohabiting/married), and schooling level (none, primary, secondary, higher). The model was fit exploring all potential main effects of socio-demographic variables and interactions with gender. A final model was obtained by model averaging all models within 4 AIC units of the top model (48). The full details of model fitting are described in the Supplemental Materials.

Using this model, we predicted an estimated degree percentile for each individual in the two networks. These predicted degree percentiles were then used to rank individuals by their estimated degree centrality and these rankings were used to prioritize testing in the targeted strategy. All other aspects of the simulations (e.g. transmission dynamics, control strategies) remained unchanged. We then compared the efficiency of the two control strategies on known and unknown network topologies, where testing was prioritized based on individuals’ true and statistically-estimated degrees, respectively.

## Data Availability

All code and data needed to reproduce the SEIR simulations will be freely available via a figshare
repository upon publication. Socio-demographic data is protected under an existing data agreement and is only available upon request.

## ACKNOWLEDGMENTS

We would like to thank the communities of Mandena and Sarahandrano for their participation in this study. We would also like to thank Jessica Metcalf for her feedback on earlier drafts of the manuscript.

## ETHICAL STATEMENT

The Institutional Review Board (IRB) at Duke University (protocol no. 2019-0560) and Malagasy Ethics Panel (137 MSNP/SG/AGMED/CERBM du 26 août 2020) approved the survey protocols used in this study. Participants in the social surveys and wearing GPS trackers provided written and verbal consent.

## DATA SHARING

All code and data needed to reproduce the SEIR simulations will be freely available via a figshare repository upon publication. Socio-demographic data is protected under an existing data agreement and is only available upon request.

## Contributors

MVE, TR, and BR conceptualized the study. All authors contributed to designing the methodologies concerning data collection and analysis. KK, CN, JM, JYR, PR, TMR, VS, and GT collected the data and MVE, TR, KK, and GT conducted analyses. AG, CN, JM, and BR provided project supervision. MVE, KK, and GT wrote the initial draft of the manuscript. All authors contributed critically to the drafts and gave final approval for publication. All authors confirm that they had full access to all the data in the study and accept responsibility for submission for publication.

## Notes

### Competing Interest Statement

The authors have declared no competing interest.

### Funding Statement

This research was funded by the Agence Nationale de la Recherche, a NIH-SSF-NIFA Ecology and Evolution of Infectious Disease Award (No. 1R01-TW011493-01), and a Duke University Provost's Collaboratory grant

### Author Declarations

The Institutional Review Board (IRB) at Duke University (protocol no. 2019-0560) and Malagasy Ethics Panel (137 MSNP/SG/AGMED/CERBM du 26 aout 2020) approved the survey protocols used in this study.

### Summary of Updates

This version of the manuscript contains a new SEIR model formulation that includes presymptomatic transmission, imperfect isolation, and waning immunity of recovered individuals. It has also been reparameterized for more recent SARS-CoV-2 variants.

