## Supplemental Materials for "Leveraging social network topology could improve the efficiency of SARS-CoV-2 epidemic control strategies in resource-limited contexts"

### Targeting SARS-CoV-2 superspreading infections could dramatically improve the efficiency of epidemic control strategies in resource-limited contexts

MV Evans*, T Ramiadantsoa, K Kauffman, C Nunn, J Moody, JY Rabezara, P Raharimalala, TM

Randriamoria, V Soarimalala,G Titcomb, A Garchitorena, B Roche

**Table of Contents**

1. Close contact network model specification for Sarahandrano
2. Model specification
3. Demographic predictors of degree centrality
4. Sensitivity analysis of intervention, biological, and network parameters
5. French summary and figures

#### 1. CLOSE CONTACT NETWORK MODEL SPECIFICATION FOR SARAHANDRANO

The close contact network for Sarahandrano was derived using the same methods as the Mandena network, as described by Kauffman et al. (1). Because this network has not yet been published, we describe the data collection and analysis methods in full here.

Participants were surveyed using the snowball sampling technique, and all survey participants were eligible to wear a GPS tracking device (iGot-U 120; Mobile Action, New Taipei City, Taiwan). The data collection occurred over the course of 3 field seasons (November to December 2020, March to May 2021, and July to September 2021). During each field season we surveyed 65, 189 and 181 people respectively and of those participants collected usable GPS data (after accounting for device malfunction and participant compliance with wearing the device) on 43, 157, and 121 people for a total of 321 individuals with GPS data, or 51360 dyads. Participants from previous seasons were not eligible to wear a GPS in subsequent seasons. GPS devices were programmed to record 6 days of the week (excluding Fridays, the day of device distribution). The mean duration of tracking device function was 5.3 ± 1.4 days during the first season, 5.2 ± 1.35 days the second season, and 5.0 ± 1.7 days the third season.

All data preparation and analyses were completed in R v. 4.1.2. The packages used: tidyverse (1.3.1), lubridate (1.8.0), igraph(1.2.11), adehabitatHR (0.4.19), sp (1.4-6), sf(1.0-6), raster (3.5-15), move (4.1.6), maptools (1.1-2), tidystringdist (0.1.4), statmod (1.4.6), ROCR (1.0-11), tweedie (2.3.3), kader (0.0.8), MuMIn (1.43.17), ergm (4.1.2 and patch *i427-gof-missing*), network (1.17.1), and reshape2 (1.4.4).

GPS data were prepared using the methods described by Kauffman et al (1). The study area was 34.8 km2 and included 98.40 percent of the recorded GPS data. Using the same 1 ha minimum convex polygon (MCP) method to identify days participants did not wear their tracking device, 2101 days of data (753523/1277092 total GPS fixes) were removed from the dataset. The observed close contact network of all individuals with a minimum of 240 simultaneous fixes contained information on 8.73% (n = 4483) of possible edges. This was used to impute the full close contact network in a two step process using an exponential random graph model (ERGM) to determine the probability an edge existed, multiplied its edge weight, as predicted by the general linear model (GLM). The possible predictors are the same as the Mandena network reported by Kauffman et al (1).

The best resulting ERGM includes the terms edges, VI 95 during the daytime and nighttime, the number of times participants named each other, if they did not name each other, only one named the other, or if both named each other (reciprocity), the distance between their houses, if they are the same or different genders, geometrically weighted edgewise-shared partner (GWESP), and geometrically weighted non-edgewise-shared partner (GWNSP). The model containing naming information was used here, and not in the first network, because the aims of our analyses would have been confounded by using the naming data. The chosen model had a sensitivity of 0.5979, specificity of 0.9502 and accuracy of 0.8700. Among the 1000 simulated networks, 5.42% (n = 2786) of edges were present in every simulation and 0.45% (n = 230) of edges were not present in any simulations.

The best resulting GLM included the cube root of the volume intersection of the home ranges (95%), the interaction between the distance between individuals' houses and if they live <25m apart, and a naming based edge of if the individuals did not name each other, one named the other, or both named each other. This model has a sensitivity of 0.8359, specificity of 0.7969 and accuracy of 0.8096 using a threshold of 0.001, which is equivalent to about one contact every other day (0.96).

#### 2. MODEL SPECIFICATION

At each time step *t*, equivalent to one day, an individual could become susceptible, exposed, infected, or recovered (Fig. S2.1). To become exposed, a susceptible individual must have been connected to an infected individual via a network edge. For each contact event, an individual's probability of becoming exposed was a function of the transmission probability $\beta$ of the infected contact. Exposed individuals became infected with a time-dependent probability $\alpha$. We assumed thirty percent (30%) of infected individuals were asymptomatic (2), and that asymptomatic individuals had a transmission rate equal to 30% of the transmission rate ($\beta$ ) of symptomatic individuals (Bi et al. 2021). All infected individuals recovered with time-dependent probability $\gamma$, and recovered individuals became susceptible with time-dependent probability ζ.

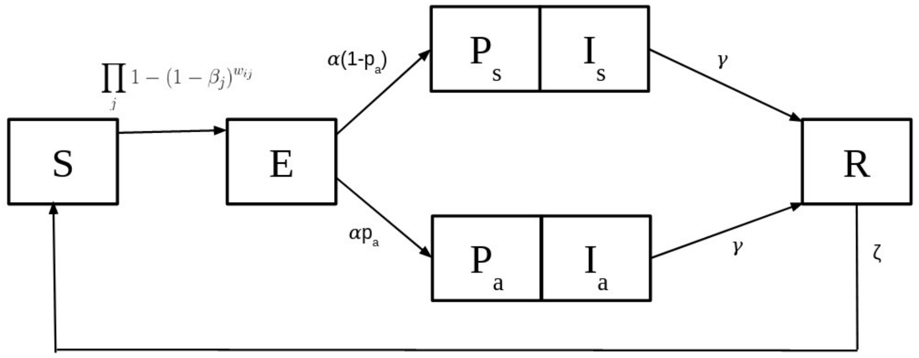


**Figure S2.1.** Compartmental diagram for SEIR model.

Each transition event was drawn from a Bernoulli distribution defined by a daily probability $\beta$, $\alpha$, $\gamma$, or ζ. Time-dependent transition rates more accurately describe SARS-CoV-2 epidemic dynamics than memoryless transition rates (3). Therefore, we relaxed the memoryless assumption of all three transition rates and assumed that they were a function of each individual's cumulative time in that state, known as non-constant hazard rates. These curves were fit so that individuals were most infectious at the beginning of their infectious period (3) (Fig. S2.3), the incubation period followed a gamma distribution with a mean of 3.1 days (*sd* 2.6), including one day of pre-symptomatic transmission (4), the mean recovery period followed a Poisson distribution with a mean of 9.67 days (5) and waning immunity followed a gamma distribution with a mean of 90 days (*sd* 20.12), parameterizing the scale of the distribution so that recovered individuals had a probability of becoming susceptible beginning 40 days post infection, with that probability increasing logistically until 150 days post-infection (Fig. S2.2).

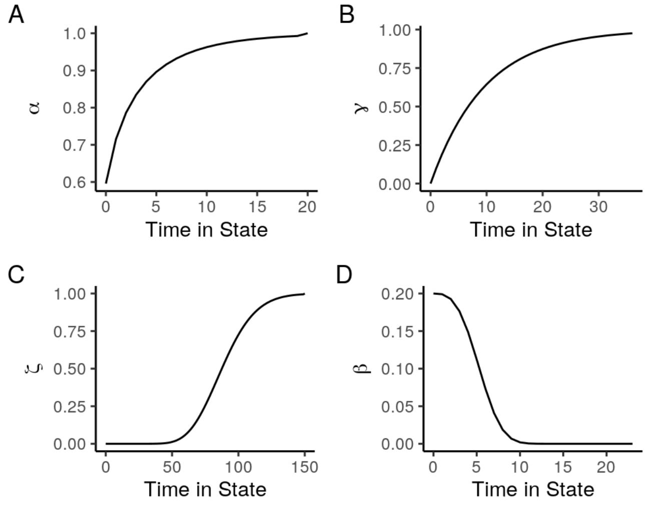


**Figure S2.2**. Time-dependent transition rates.

The transmission rate, $\beta$, was chosen to approximate an epidemic that infects around 75% of the susceptible population at the end of the outbreak and resulted in a realistic reproduction number at the early stages of the epidemic, as detailed below. We calculated the average effective reproductive number during the first month of the epidemic following Cori et al. (7), assuming a mean serial interval of 4.8 days (4). We estimated the effective reproduction number on a network of 10,000 nodes with degree and edge-weight distributions identical to those of the field-derived social network from Mandena, to approximate spread on a network without density-dependent effects, which were strong on our relatively small networks. Because our model was stochastic, we simulated 100 epidemics on this network, exploring a range of $\beta$ from 0.02-0.30 in steps of 0.02. The effective reproduction number is somewhat dependent on characteristics of the population where it is measured, resulting in a range of estimated effective reproduction numbers for COVID-19 in the literature. We chose to model epidemics with high transmission because vaccination rates are relatively low in Madagascar. We then chose the $\beta$ value that resulted in a mean estimated effective reproduction number during exponential growth assuming no immune evasion for an epidemic of SARS-CoV-2 Omicron variant estimated to be 24 in South Africa in December 2021(8). The final value of $\beta$ was 0.2. This resulted resulting in a mean estimated effective reproduction number during exponential growth of 19.60 and 28.52 and on day 5 of 8.99 and 12.48 on the Mandena and Sarahandrano networks, respectively. Because of the uncertainty regarding the transmission rate of COVID-19 in a novel population, we also studied the sensitivity of our analysis to changes in this transmission rate (described in section 4). We used this value of $\beta$ as the maximum transmission rate when simulating SEIR models on weighted simulated networks. An infected individual's transmission rate decreased exponentially from this maximum transmission rate as a function of the time spent in the infectious class (Fig. S2.2).

The field-derived contact network contained weighted edges, where weight was dependent on the number of 3 minute-periods in close-contact between nodes over one week. We defined an edge as having a weight equal to at least one hour in contact over a week-long period. In order to create a time-integrated network, we rescaled this weight so that a value of 1 corresponded to 24 hours in contact over the course of the week. Probability of transmission between two nodes *i* and *j*, given one node was infected and the other susceptible, was then dependent on the weight of this edge via the following equation :

$$P(i_{S\to E}) = \prod_j 1 - (1 - \beta_j)^{w_{ij}}$$

Where $\beta_j$ is the base transmission rate for infected individual *j* and $w_{ij}$ is the weight between the two nodes.

Each SEIR simulation was initiated with 2 exposed individuals at the first day of the latent period. The number of susceptible, exposed, infected (asymptomatic and symptomatic), and recovered individuals were recorded at each time step. Epidemics were simulated until no exposed, presymptomatic, infected, or isolated individuals remained, which we define as the full duration of the epidemic. The length of each individual simulation therefore depended on each simulation's initial exposed individuals and epidemic parameters. To ensure simulations included the full duration of the epidemic, we simulated for a maximum of 300 days, which was longer than all simulated epidemics.

#### 3. DEMOGRAPHIC PREDICTORS OF DEGREE CENTRALITY

Spatially-explicit contact data can enable interventions that interrupt disease transmission. However, there are numerous logistical challenges in collecting these high-resolution data, especially in rural communities. Therefore, we explored whether simple, easy-to-measure demographic variables could predict degree centrality in a contact network, and therefore, whether they could be useful in generating a ranked list of priority individuals for testing and treatment. Here, we describe how we fit a predictive model of degree centrality that was used to create the rankings used for testing prioritization when applying superspreader-prioritized strategies to networks with unknown topologies.

*Methods*

Because we were interested in building a predictive model that generalized over both networks, we combined degree data from the two villages of Mandena and Sarahandrano. To control for different network sizes (and thus, mean degree centrality), we calculated each individual's degree percentile in their respective network. We then conducted beta regression on percentile, using predictors for age, household size, marital status (single vs. cohabiting/married), and schooling level (none, primary, secondary, higher). Because men and women may differ in their effect size and direction for each of these variables, we also included the interaction between each of these predictors and gender. We also included the effect of the village in the full model.

Several data cleaning and imputation steps were needed to fit this model.

1) For the survey question relating to marital status, individuals in the two different villages were presented with a different set of response options. To simplify and align the data for the two villages, we assigned the categories of "Widowed" and "Never married/Never lived together" to be "Single". We considered "Living together", "Cohabiting", and "Married" to be synonymous because few individuals in either village were legally married, but were in relationships that were otherwise identical to marriage (e.g. living and raising children together). These individuals were described as "Cohabiting".
2) One individual in the Mandena network did not provide household size data. Given that this person listed their marital status as single and with two children, we assigned their household size to be 3.
3) One individual in each network did not provide age data. Therefore, we predicted age for the Mandena individual by building a simple linear regression model of age using household size, school level, marital status, and land size as predictors. The Sarahandrano individual did not provide data on land size, so a model without this predictor was used to estimate age.
4) One individual in the Sarahandrano network did not provide schooling data. Therefore, we used data on age and gender to determine the most probable school level.

After checking full model diagnostics, we generated and compared a list of all possible model subsets. We performed model averaging using the top models ($\Delta$ AIC < 4) to generate a final model (Figure S3.1). We then used the final model to predict each individual's degree percentile and created a ranked degree percentile list to inform testing and treatment order.

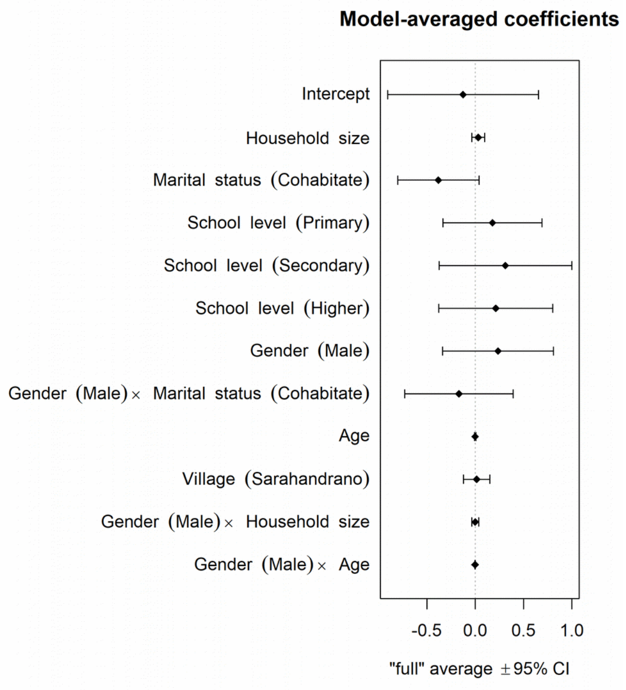


**Figure S3.1.** Full model coefficient estimates from the averaged model. Points represent mean and error bars represent 95% CI.

*Results*

We found that demographic variables were generally poor predictors of degree centrality in the contact network. The only predictor in all models was marital status, in which cohabiting individuals tended to have lower degree centrality than single individuals. However, we also note that this effect was much stronger for individuals in the Sarahandrano network than the Mandena network, indicating that there may be important social differences between these two locations (Fig. S3.2). Aside from marital status, gender, household size, and schooling level were included in nearly half of the top models, and age and village ID were each found in approximately a third of the top models (Table S3.1). Despite the relatively low $R^2$ value (0.03) for the averaged model, we found that the predicted top 10 individuals from each network had an average degree that was approximately 1.5-2 times as high as the predicted bottom 10 individuals (Figure S3.3).

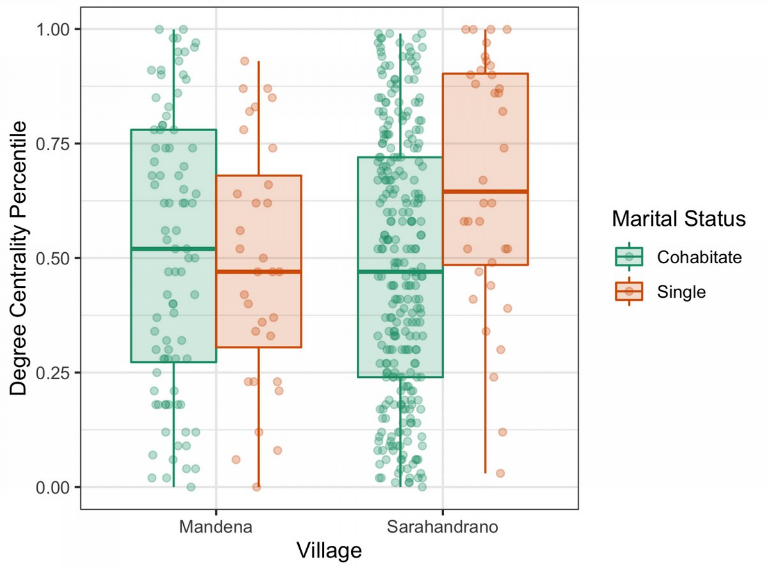

**Figure S3.2.** Degree centrality percentile by marital status and village. Note that single individuals in Sarahandrano are among the most well-connected people in the contact network, while there is little variation between the two in the Mandena network.

**Table S3.1.** Variable importance and proportion of models they occur in for the demographic predictors found in the top models (delta AIC<4) of degree centrality.

| | Marital status | Gender | Household size | School level | Gender: Marital status | Age | Village | Gender: Household size | Age: Gender |
|---|---|---|---|---|---|---|---|---|---|
| **Sum of weights** | 1 | 0.69 | 0.67 | 0.56 | 0.37 | 0.31 | 0.24 | 0.09 | 0.02 |
| **Proportion models** | 1 | 0.72 | 0.64 | 0.54 | 0.38 | 0.42 | 0.36 | 0.14 | 0.04 |

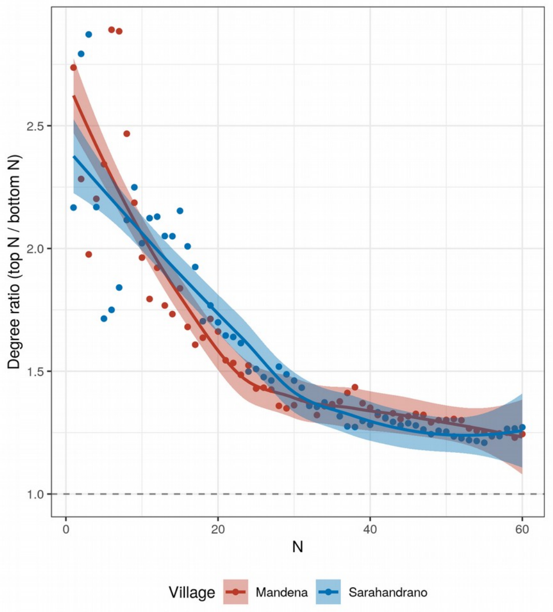


**Figure S3.3.** The averaged model differentiated individuals with high versus low degree centrality such that the top 20 individuals had at least 1.5 times the degree centrality of the bottom 20 individuals. The x-axis represents different window sizes (N) for comparing the top N individuals to the bottom N individuals. The smoothed line is a loess curve fit to the data with 95% confidence intervals. The horizontal dashed line represents no differentiation. Points are colored by the network they describe.

#### 4. SENSITIVITY ANALYSIS OF INTERVENTION, BIOLOGICAL, AND NETWORK PARAMETERS

*Methods*

We used sensitivity analyses to assess the robustness of our results to chosen parameters pertaining to the testing intervention (start date and testing ascertainment), transmission biology (transmission rate), and the social network topology (size and assortativity). For each parameter, we held all other parameters equal to in the primary model, and explored a range of values for the focal parameter (Table S4.1). For intervention and biological parameters, simulations were run on the field-derived social contact network from Mandena, with 100 simulations of each control strategy for each parameter value. For network size, we simulated 10 networks of each size and simulated 10 epidemics on each for each control strategy, resulting in 100 simulations for each control strategy for each network size. For network assortativity, we created contact networks of different levels of assortativity by rewiring the Mandena network following Xulvi-Brunet and Sokolov (9), which alters assortativity while preserving a network's degree distribution. We simulated five networks with varying levels of assortativity (Table S4.1), and simulated 100 epidemics on each network for each control strategy.

**Table S4.1.** Range and resolution of parameters used in sensitivity analyses.

| | Range | Resolution |
|---|---|---|
| **Intervention** | | |
| Testing Start Date (days) | 0 - 24 | 3 |
| Testing Ascertainment (probability of testing identified individual) | 0.1 - 1 | 0.1 |
| **Biological** | | |
| Transmission Rate | 0.05 - 0.3 | 0.05 |
| **Network Characteristics** | | |
| Size | 200 - 10,000 | Specific Values: 200, 500, 1000, 2500, 5000, 10000 |
| Assortativity | -0.14 - 0.22 | Specific values: -0.14, -0.09, -0.07, 0.15, 0.22 |

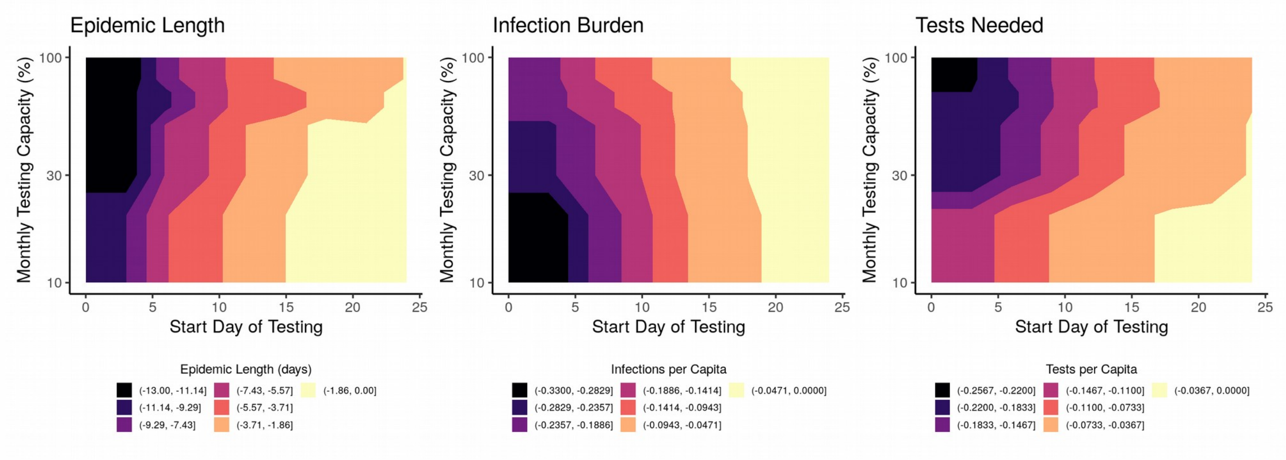


**Figure S4.1.** Difference in mean efficiency between testing randomly or prioritizing testing according to degree centrality across different testing start dates. Color represents the mean of 100 simulations, with darker colors representing when targeted testing was most effective.

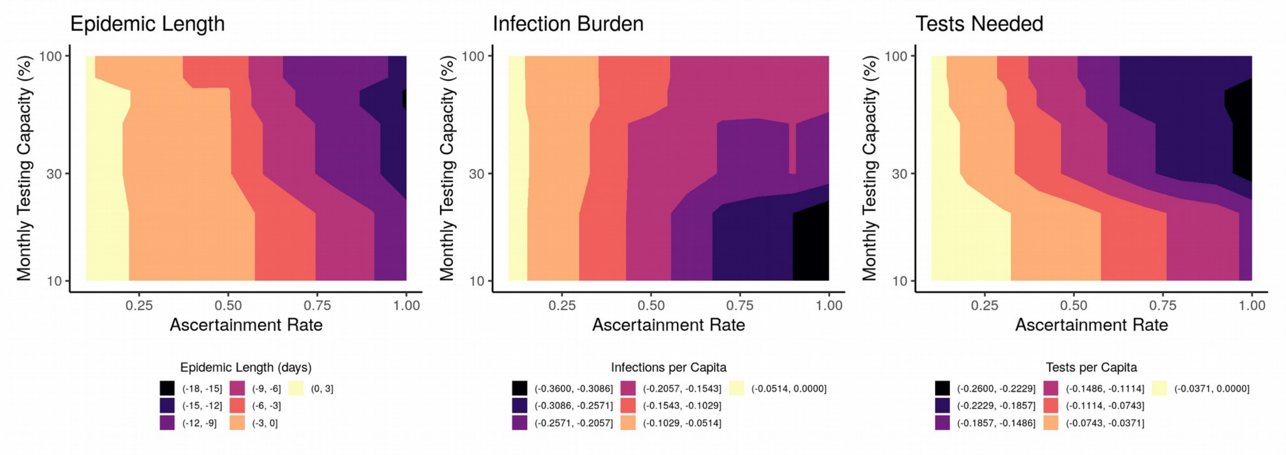


**Figure S4.2.** Difference in mean efficiency between testing randomly or prioritizing testing according to degree centrality across different ascertainment rates. Color represents the mean of 100 simulations, with darker colors representing when targeted testing was most effective.

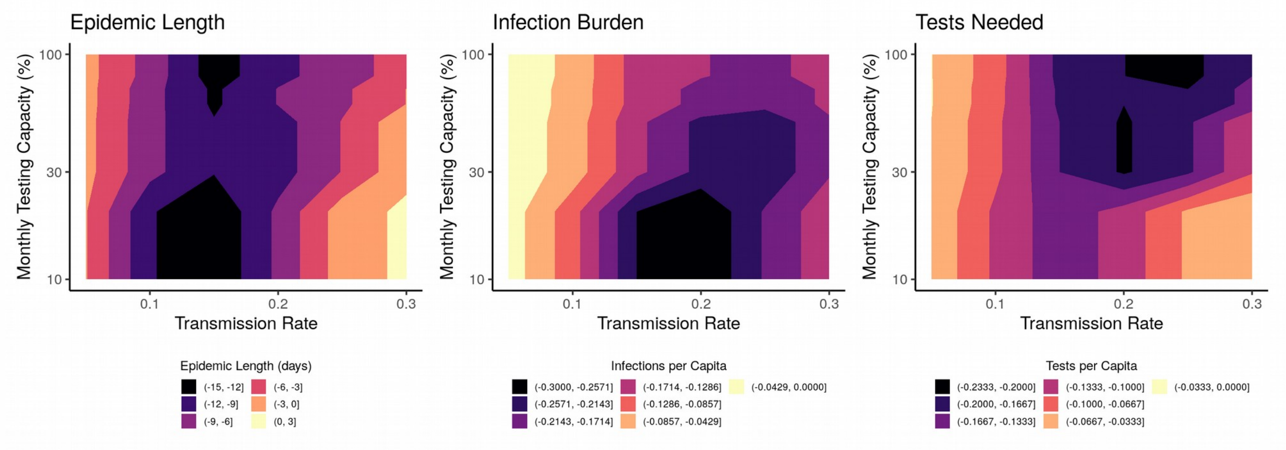


**Figure S4.3.** Difference in mean efficiency between testing randomly or prioritizing testing according to degree centrality across different transmission rates. Color represents the mean of 100 simulations, with darker colors representing when targeted testing was most effective.

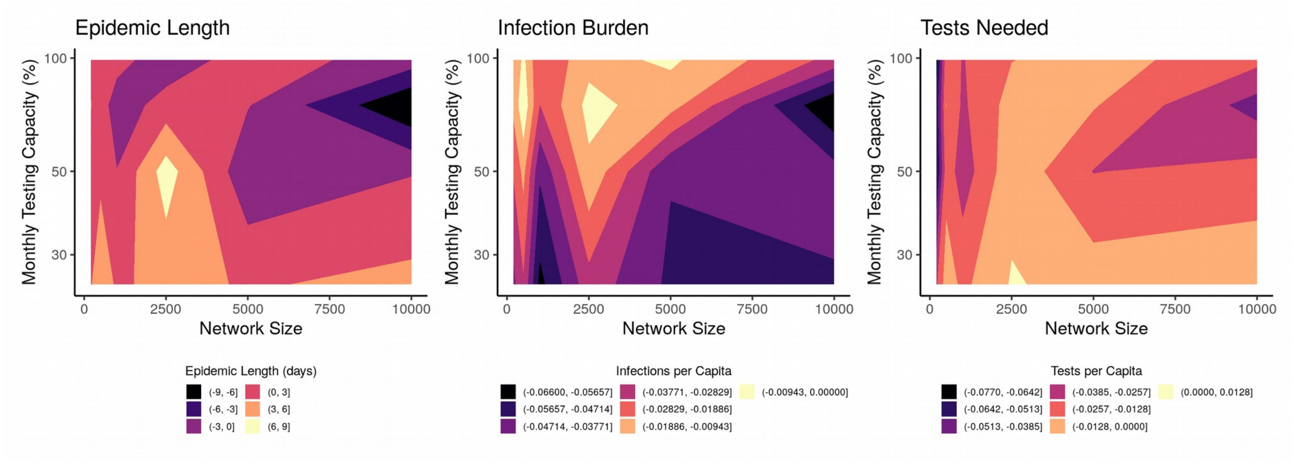


**Figure S4.4.** Difference in mean efficiency between testing randomly or prioritizing testing according to degree centrality across different network sizes. Color represents the mean of 100 simulations, with darker colors representing when targeted testing was most effective.

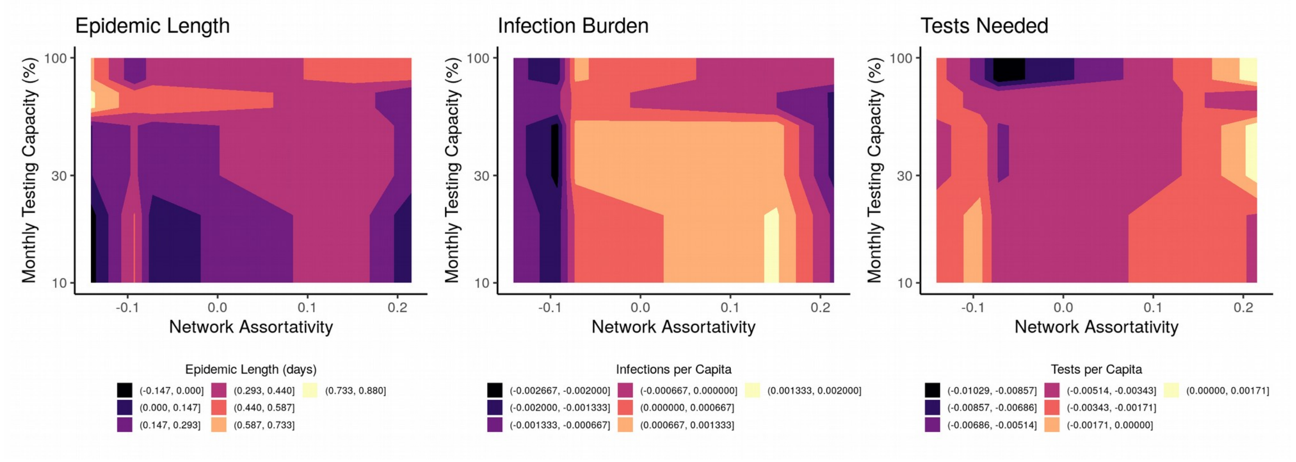


**Figure S4.5.** Difference in mean efficiency between testing randomly or prioritizing testing according to degree centrality across networks with varying assortativity values. Color represents the mean of 100 simulations, with darker colors representing when targeted testing was most effective.

#### 5. FRENCH SUMMARY & FIGURES

Following medrXiv policy, we are unable to provide the French translation of the summary and figures. Please contact the corresponding author (mv.evans.phd@gmail) for access to these documents.
